# Logistic regression with machine learning sheds light on the problematic sexual behavior phenotype

**DOI:** 10.1101/2022.01.31.21267382

**Authors:** Shui Jiang, Keanna Wallace, Esther Yang, Leslie Roper, Garima Aryal, Dawon Lee, Rohit J Lodhi, Rick Isenberg, Bradley Green, David Wishart, Katherine J Aitchison

**Affiliations:** Department of Medical Genetics, Faculty of Medicine and Dentistry, College of Health Sciences, University of Alberta, Canada; Department of Psychiatry, Faculty of Medicine and Dentistry, College of Health Sciences, University of Alberta, Canada; Department of Emergency Medicine, Faculty of Medicine and Dentistry, College of Health Sciences, University of Alberta, Canada; Department of Medicine, Faculty of Medicine and Dentistry, College of Health Sciences, University of Alberta, Canada; Department of Psychiatry, University of Western Ontario, Canada; Psychological Counselling Services, Ltd., Scottsdale, USA; Department of Psychology and Counseling, University of Texas at Tyler, USA; Department of Biological Sciences, Faculty of Science, College of Natural and Applied Sciences, University of Alberta, Canada; Neuroscience and Mental Health Institute, University of Alberta, Canada; Division of Clinical Sciences, Psychiatry Section, Northern Ontario School of Medicine

**Author notes:** Corresponding author: (KJA).

## Abstract

**Objectives:** There has been a longstanding debate about whether the mechanisms involved in problematic sexual behavior (PSB) are similar to those observed in addictive disorders, or related to impulse control or to compulsivity. The aim of this report was to contribute to this debate by investigating the association between PSB, addictive disorders (internet addiction, compulsive buying), measures associated with the construct known as reward deficiency (RDS), and obsessive-compulsive disorder (OCD) in young adults in post-secondary education.

**Methods:** A Canadian university Office of the Registrar invited 68,846 eligible students and postdoctoral fellows. Out of 4710 expressing interest in participating, 3359 completed on-line questionnaires, and 1801 completed the Mini-International Neuropsychiatric Interview (MINI). PSB was measured by combining those screening positive (≥6) on the Sexual Addiction Screening Test-Revised (SAST-R) Core with those self-reporting PSB. Current mental health condition(s) and childhood trauma were measured by self-report. OCD was assessed by a combination of self-report and MINI data.

**Results:** 12.18% (407/3341) of participants screened positive on the SAST-R Core. On logistic regression, OCD, attention deficit, internet addiction, a family history of PSB, childhood trauma, compulsive buying and male gender were associated with PSB. On multiple correspondence analysis (MCA), OCD appeared to cluster separately from the other measures, and the pattern of data differed by gender.

**Conclusions:** Factors that have previously been associated with RDS and OCD are both associated with increased odds of PSB. The factors associated with RDS appear to contribute to a separate data cluster from OCD, and to lie closer to PSB.

## INTRODUCTION

A theory of hypersexuality with dependence was first proposed by Orford in 1978.^1^ The term sex addiction was used by Carnes in 1983, 1990, and 1991^2^ and by Goodman in 1998.^3^ Continuation of a sexual behavior despite adverse consequences and/or distress caused or worsened by the sexual behavior has been a consistent theme in the literature,^1,3,4^ despite differences of opinion as to whether the condition should be classified as an addiction or as an impulse control disorder.

Diagnostic criteria proposed by Carnes in 2005^4^ led to sex addiction being included in the Diagnostic and Statistical Manual of Mental Disorders (DSM)-III-R (302.87). However, despite draft proposed criteria for the inclusion of sex addiction and internet addiction in DSM-5,^5^ the only behavioral addiction currently included in the main body of DSM-5 (American Psychiatric Association 2013) is gambling disorder (312.31), which was first included in DSM as an impulse control disorder.^6^ In the International Statistical Classification of Diseases (ICD), compulsive sexual behavior disorder was proposed for inclusion as an impulse control disorder in 2014,^7^ which occurred in 2018.^8^ However, the scientific discussion about whether or not compulsive sexual behavior disorder could constitute a manifestation of a behavioral addiction was acknowledged,^9^ and it was predicted (as in the case of problematic gambling),^6^ that understanding would evolve as research elucidated the phenomenology and neurobiological underpinnings of the condition.

In this article, the term problematic sexual behavior (PSB) is used because it focuses on the behavior rather than on the potentially heterogeneous etiology and pathology.^10^ PSB is characterized by repetitive sexual behaviors and it is associated with uncontrolled sexual urges/impulses and distress,^11^ and social and functional impairments.^8,11^ It has been estimated that the prevalence of PSB is 3-6% in adults,^4,12-14^ with higher frequencies in specific populations, including those in post-secondary education. Reid (2011) reported that 19% of college men met criteria for hypersexuality,^15^ while Giordano and Cecil (2014) found 11.1% of men and women in an undergraduate sample met these criteria.^16^ Cashwell et al. (2015) reported that out of 379 undergraduates, 21.2% of men and 6.7% of women screened positive, i.e., scored in a range indicating that they should be offered further assessment for sex addiction.^17^

Various vulnerability/risk factors have been associated with PSB, including those previously associated with the construct known as reward deficiency syndrome (RDS) (substance^18^ and non-substance use disorders,^19^ ADHD,^20^ and personality disorder),^21^ as well as other psychiatric disorders,^22^ and childhood trauma.^23^ In terms of psychiatric disorders, owing to the classification of PSB within ICD-10 as an impulse control disorder, we specifically included obsessive-compulsive disorder. Mick and Hollander (2006) hypothesized that impulsivity initiated the early stage of PSB, with compulsivity being involved in repetitive behaviors and hence in the maintenance of PSB.^24^ The magnitude of the contribution of the various different postulated mechanisms, and whether PSB encompasses various syndromes with different etiologies is not at present known. Moreover, systematic data on the prevalence of PSB and associated sociodemographic factors across diverse populations, including non-treatment seekers remain to be provided.^8^

In light of the above, this paper aims to further elucidate the prevalence, sociodemographic features, phenomenology, and neurobiological underpinnings of the construct known as PSB in a diverse sample of adults in post-secondary education. We hypothesized that in this sample, firstly, screening positive for factors previously associated with RDS (such as internet addiction, compulsive buying, nicotine dependence, and pathological gambling) and secondly, having OCD and childhood trauma would be associated with increased odds of PSB.

## METHODS

The study inclusion criteria were as follows: at least 18 years of age, undergraduates, graduates, postdoctoral fellows, and recently convocated students registered in at least one course in the preceding year, except for those who had completely withdrawn after registration without any reasons to not return in the next academic term or not be on campus (such as suspension), and being able to answer in English. A Canadian university Office of the Registrar invited 68,846 eligible students and postdoctoral fellows by email. Students interested in participating emailed the study team. These students were then sent an email invitation to review the participant information and complete consent on-line, followed by the screening measures (hosted by the Qualtrics platform), with a subsequent email inviting them to complete the Mini-International Neuropsychiatric Interview (MINI) version 7.0.2 on-line.

### Measures

The Sexual Addiction Screening Test-Revised (SAST-R)^25^ is a 45-item screening tool comprising several subscales designed to detect potentially problematic sexual behavior. Each question is answered in a binary manner (yes/no=0/1). Sexual activities deemed to be problematic by participants in the past 30 days were also separately assessed by asking the following question: “Within the last 30 days, how often have you participated in activities of a sexual nature that you would regard as problematic?”

Measures used to screen for internet addiction, compulsive buying, personality disorders, ADHD, nicotine dependence, and pathological gambling were: the Internet Addiction Test (IAT),^26^ the Richmond Compulsive Buying Scale (RCBS),^27^ the 8-item Standardized Assessment of Personality–Abbreviated Scale as a Self-Administered Screening Test (SA-SAPAS),^28^ the full Adult ADHD Self-Report Scale (ASRS-v1.1),^29^ the Fagerström Test for Nicotine Dependence (FTND),^30^ and the DSM-V Pathological Gambling Diagnostic Form (PGDF),^31^ respectively. Gender, sexual orientation, a family history of domestic violence or of sex addiction, and having received a diagnosis of OCD, or any other mental health condition(s), childhood physical, emotional/verbal, or sexual trauma were additionally collected by self-report. The MINI was used to output psychiatric disorders by DSM-5 criteria.

## Data analysis

### Demographic data and logistic regression

Data were analyzed by STATA (Stata/SE 16) and R (Version 3.6.3) after dropping the missing data. The PSB variable was created by combining those screening positive on the SAST-R Core with those endorsing monthly self-reported PSB, after subtracting the two SAST-R Core questions referred to below. Previous or current physical/verbal, emotional/sexual childhood trauma and a family history of domestic violence (11.77% 387/3288) were regrouped as total childhood trauma due to the relatively low rates of endorsement for the former group (physical trauma: 2.98%, 99/3322; emotional trauma: 6.87%, 232/3322; and sexual trauma: 3.31%, 110/3212). The SAST-R Core question 1 (which enquires about a history of sexual abuse in childhood or adolescence, SASTC1) was combined with total childhood trauma. Self-reported a family history of sex addiction (“yes”/ “no”) data were combined with SAST-R Core question 2 (which enquires about parental trouble with sexual behavior, SASTC2) to create a new family history of PSB variable. IAT was regrouped as “yes” (N=1174, combining mild, moderate, and severe groups) and “no” (N=2160, the “normal” group).

Differences in data distributions between those with and without PSB were assessed by Pearson’s χ^2^ test for categorical variables and by non-parametric testing (Mann-Whitney U test) for linear non-normally distributed variables. For correlational analyses, tetrachoric (for binary variables) and Kendall’s τ (for categorical variables) correlations were utilized. SA-SAPAS, OCD, ASRS, IAT, childhood trauma, a family history of PSB and gender were used as covariates in the regression analyses. All reported *P* values are unadjusted for multiple testing. Multiple correspondence analysis (MCA) was used for data clustering.

### Machine learning approaches

The “Haven” and “dplyr” packages were used for data input. The “forcats” and “creditmodel” packages were used for recoding and splitting the dataset (into 80% training and 20% testing), respectively. The Synthetic Minority Oversampling Technique (SMOTE) from the “Caret” package was used to address data imbalance in the training set.^32^ Leave-one-out cross-validation (LOOCV) in the “Caret” package was used for resampling.^33^ Logistic regression (two-class summary) in the “caret” package was used for regression. Accuracy, area under the receiver operating characteristic (ROC) curve, sensitivity, specificity, and F score were used to evaluate the models. The “pROC” package was used for the area under the ROC curve.

### Multiple correspondence analysis (MCA)

Multiple correspondence analysis was conducted on OCD, RCBS, SA-SAPAS, ASRS, IAT, and childhood trauma, stratifying by gender (women N=2084 and men N=1012). As a family history of PSB was correlated with the combined childhood trauma variable (tetrachoric ρ=0.69, *P*<0.001, N=3019, Figure 1, Supplemental Digital Content, Table 3), these variables were combined as a supplementary childhood trauma-related variable. The “FactoMineR,” “factoextra,” and “GDAtools” packages in R were used for analysis and data visualization.

**Figure 1.**
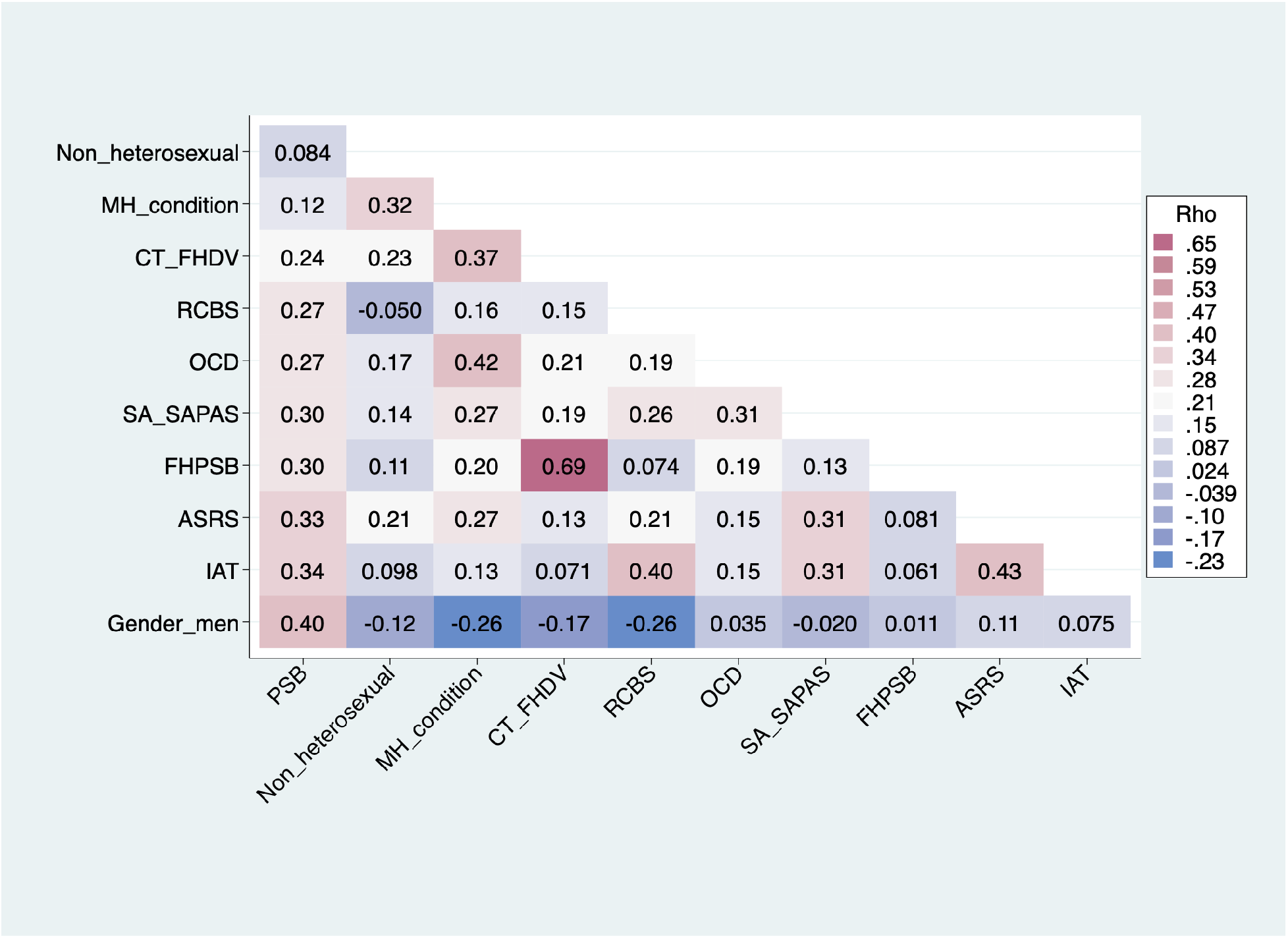
Correlation matrix for problematic sexual behavior (PSB) with demographic and clinical variables. *P* values are provided in Supplemental Digital Content, Table 3. MH_condition: mental health condition; CT_FHDV: childhood trauma and a family history of domestic violence (including physical, verbal/emotional and sexual childhood trauma and SAST-R Core question 1); RCBS: Richmond Compulsive Buying Scale; OCD: obsessive-compulsive disorder (including self-reported and MINI-diagnosis); SA-SAPAS: Standardized Assessment of Personality–Abbreviated Scale as a Self-Administered Screening Test; FHPSB: a family history of problematic sexual behavior (including a family history of sexual addiction and SAST-R Core question 2); ASRS: the Adult ADHD Self-Report Scale (v1.1); IAT: the Internet Addiction Test

## RESULTS

Out of 4710 interested in participating, 237 withdrew from the study, 3359 completed online questionnaires using the Qualtrics platform, and 1801 completed the MINI (Supplemental Digital Content, Figure 1).

### Demographic data

The mean score on the SAST-R Core Scale among all participants was 2.23 (SD=2.73), with 12.18% (407/3341) of participants scoring at least six (the threshold for screening positive in the general population.^25^ Of the 20 items composing the SAST-R Core questions, item 11 (Do you hide some of your sexual behavior from others?) was the most endorsed (1234/3341, endorsement rate: 36.78%), while item 9 (“Are any of your sexual activities against law?”) was the least endorsed (35/3342; endorsement rate: 1.05%) (Supplemental Digital Content, Table 1). The frequency of participants included in the PSB variable was 16.53% (532/3219). There was a significant difference in the distribution of the following variables by those with and without PSB: gender, sexual orientation, OCD, RCBS, a family history of PSB, SA-SAPAS, current mental health condition(s), childhood trauma, ASRS, and IAT (Table 1, and Supplemental Digital Content, Table 2).

**Table 1.** Distribution of demographic and clinical variables by problematic sexual behavior (PSB) group.

| Categories |  | PSB “yes”<br>532/3219<br>(16.53%) | PSB “no”<br>2687/3219<br>(83.47%) | Total<br>number | P value |
| --- | --- | --- | --- | --- | --- |
| SAST-R Core score | | 6.58±3.22<br>(N=532) | 1.38±1.56<br>(N=2687) | 3219 | $P<0.001^a$ |
| Age | | 22.32±5.13<br>(N=532) | 22.34±5.05<br>(N=2686) | 3218 | $P=0.81^a$ |
| Gender | Women | 232 (10.82%) | 1913 (89.18%) | 2145 | $P<0.001^b$ |
|  | Men | 290 (28.27%) | 736 (71.73%) | 1026 |  |
|  | Total | 522 (16.46%) | 2649 (83.54%) | 3171 |  |
| Sexual<br>Orientation | Heterosexual | 420 (15.85%) | 2230 (84.15%) | 2650 | $P=0.034^b$ |
|  | Non-heterosexual | 110 (19.50%) | 454 (80.50%) | 564 |  |
|  | Total | 530 (16.49%) | 2684 (83.51%) | 3214 |  |
| OCD | No | 490 (15.88%) | 2596 (84.12%) | 3086 | $P<0.001^b$ |
|  | Yes | 40 (33.33%) | 80 (66.67%) | 120 |  |
|  | Total | 530 (16.53%) | 2676 (83.47%) | 3206 |  |
| RCBS | No | 471 (15.54%) | 2559 (84.46%) | 3030 | $P<0.001^b$ |
|  | Yes | 61 (32.28%) | 128 (67.72%) | 189 |  |
|  | Total | 532 (16.53%) | 2687 (83.47%) | 3219 |  |
| **FHPSB** | **No** | 453 (15.38%) | 2492 (84.62%) | 2945 | *P*<0.001<sup>b</sup> |
|  | **Yes** | 71 (34.63%) | 134 (65.37%) | 205 |  |
|  | **Total** | 524 (16.63%) | 2626 (83.37%) | 3150 |  |
| **SA-SAPAS** | **No** | 425 (14.76%) | 2454 (85.24%) | 2879 | *P*<0.001<sup>b</sup> |
|  | **Yes** | 107 (31.47%) | 233 (68.53%) | 340 |  |
|  | **Total** | 532 (16.53%) | 2687 (83.47%) | 3219 |  |
| **Mental health condition(s)** | **No** | 399 (15.35%) | 2200 (84.65%) | 2599 | *P*<0.001<sup>b</sup> |
|  | **Yes** | 131 (21.87%) | 468 (78.13%) | 599 |  |
|  | **Total** | 530 (16.57%) | 2668 (83.43%) | 3198 |  |
| **Childhood trauma** | **No** | 339 (13.94%) | 2092 (86.06%) | 2431 | *P*<0.001<sup>b</sup> |
|  | **Yes** | 185 (25.45%) | 542 (74.55%) | 727 |  |
|  | **Total** | 524 (16.59%) | 2634 (83.41%) | 3158 |  |
| **ASRS** | **No** | 321 (12.97%) | 2153 (87.03%) | 2474 | *P*<0.001<sup>b</sup> |
|  | **Yes** | 211 (28.32%) | 534 (71.68%) | 745 |  |
|  | **Total** | 532 (16.53%) | 2687 (83.47%) | 3219 |  |
| **IAT** | **Normal (No)** | 241 (11.50%) | 1855 (88.50%) | 2096 | *P*<0.001<sup>b</sup> |
|  | **Mild, moderate or severe (Yes)** | 291 (25.91%) | 832 (74.09%) | 1123 |  |
|  | **Total** | 532 (16.53%) | 2687 (83.47%) | 3219 |  |

<sup>a</sup>Mann-Whitney U test
<sup>b</sup>Pearson's $\chi^2$ test
OCD: obsessive-compulsive disorder (including self-reported and MINI-diagnosis); RCBS: Richmond Compulsive Buying Scale; FHPSB: a family history of problematic sexual behavior (including a family history of sex addiction and SAST Core question 2); SA-SAPAS: Standardized Assessment of Personality–Abbreviated Scale as a Self-Administered Screening
Test; Childhood trauma: childhood trauma (including physical, verbal/emotional and sexual childhood trauma and SAST Core question 1) and a family history of domestic violence; ASRS: the Adult ADHD Self-Report Scale (v1.1); IAT: the Internet Addiction Test

Correlational analyses in the whole dataset after dropping the missing values showed that PSB was correlated with non-heterosexual sexual orientation (baseline: heterosexual), current mental health condition(s), childhood trauma, RCBS, OCD, SA-SAPAS, a family history of PSB, ASRS, IAT and gender (baseline: women) (Figure 1, Supplemental Digital Content, Table 3). Interestingly, a family history of PSB was shown to be strongly correlated with childhood trauma in our dataset (tetrachoric ρ=0.69, *P*<0.001, N=3019), and both were correlated with PSB. For example, of those responding “yes” to a family history of sex addiction, 32.65% (16/49) screened positive on the SAST-R Core. Our data also showed that: (A) PSB was correlated with unadjusted SAST-R Core (tetrachoric ρ=0.97, *P*<0.001, N=3219) and self-reported monthly PSB (tetrachoric ρ=0.70, *P*<0.001, N=3271), (B) self-reported OCD was correlated with OCD MINI diagnosis (tetrachoric ρ=0.47, *P*<0.001, N=1785) (Supplemental Digital Content, Table 4).

## Regression models

### Logistic regression

Outliers were removed from the logistic regression (N=47, with 8 categorized as “yes” to having PSB, Supplemental Digital Content, Figure 2) if their Pearson standardized residual had an absolute value of more than 2,^34^ or Hosmer-Lemeshow 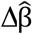 more than four.^35^ After outlier removal as above, no further outliers needed to be removed on the basis of leverage values (a measure of the distance between independent variable observations). The following variables were associated with PSB (pseudo *R*^*2*^=0.16): OCD (*P*=0.015), ASRS (*P*<0.001), IAT (*P*<0.001), a family history of PSB (*P*<0.001), SA-SAPAS (*P*<0.001), childhood trauma (*P*<0.001), RCBS (*P*<0.001) and gender (men) (*P*<0.001) (Figure 2).

**Figure 2.**
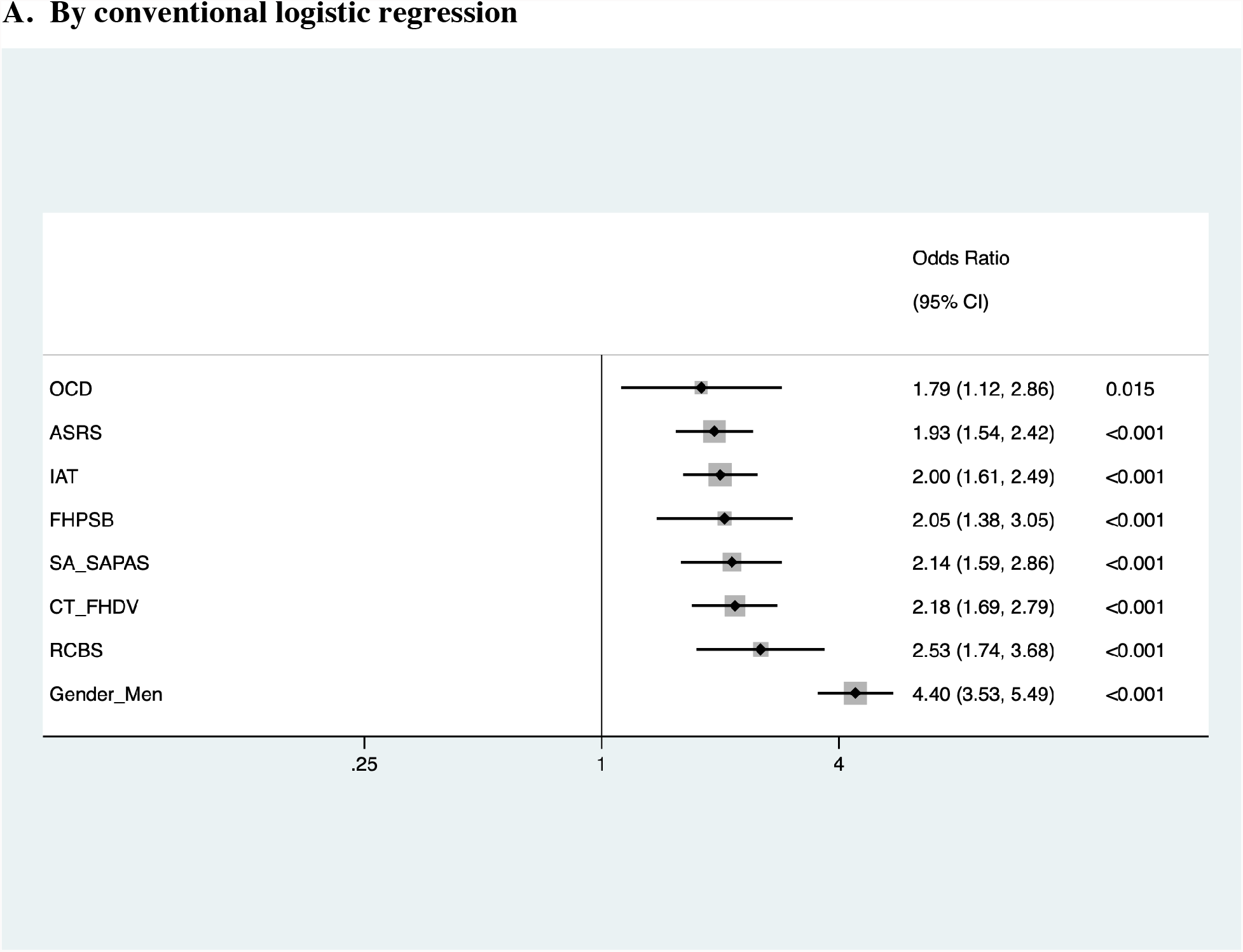
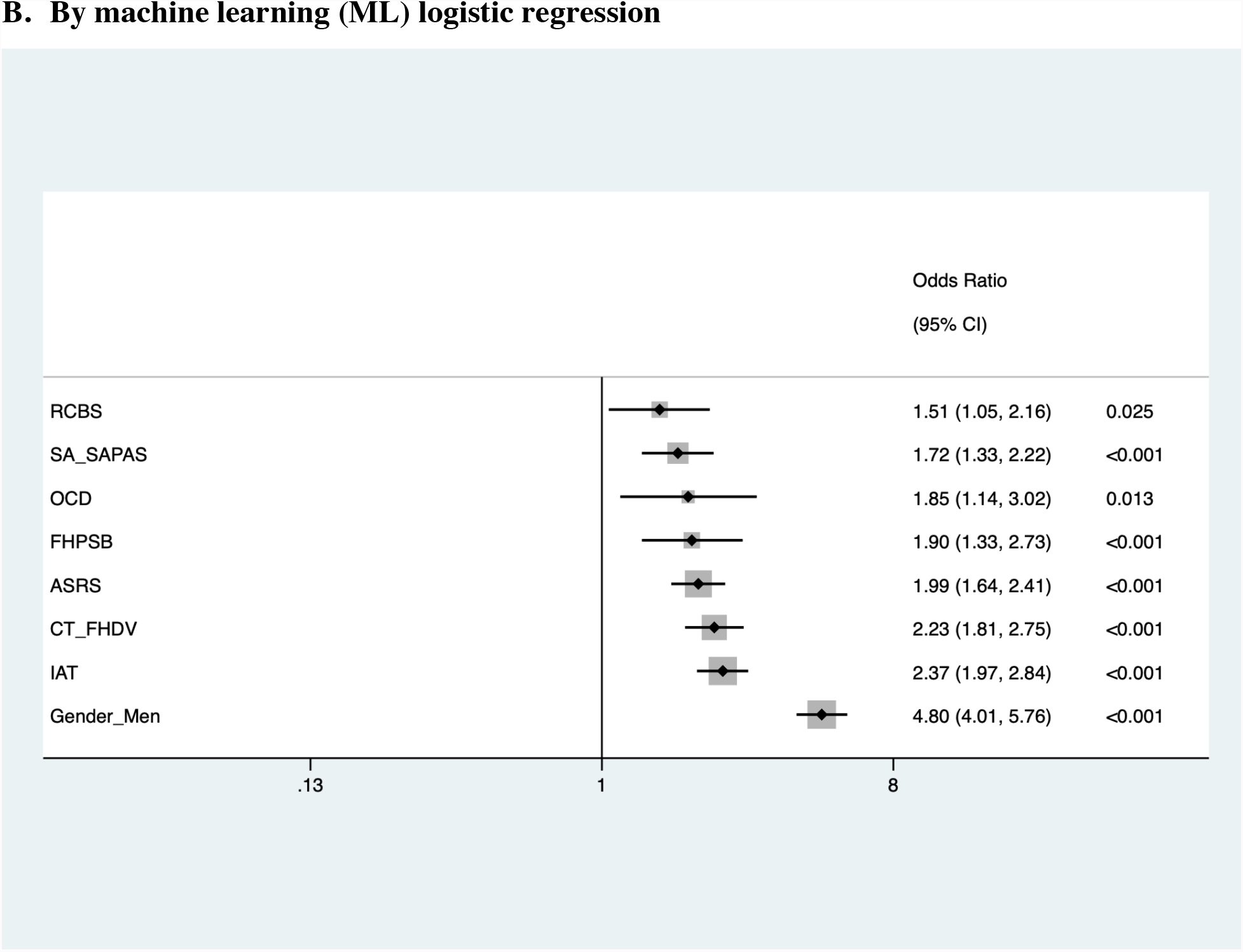
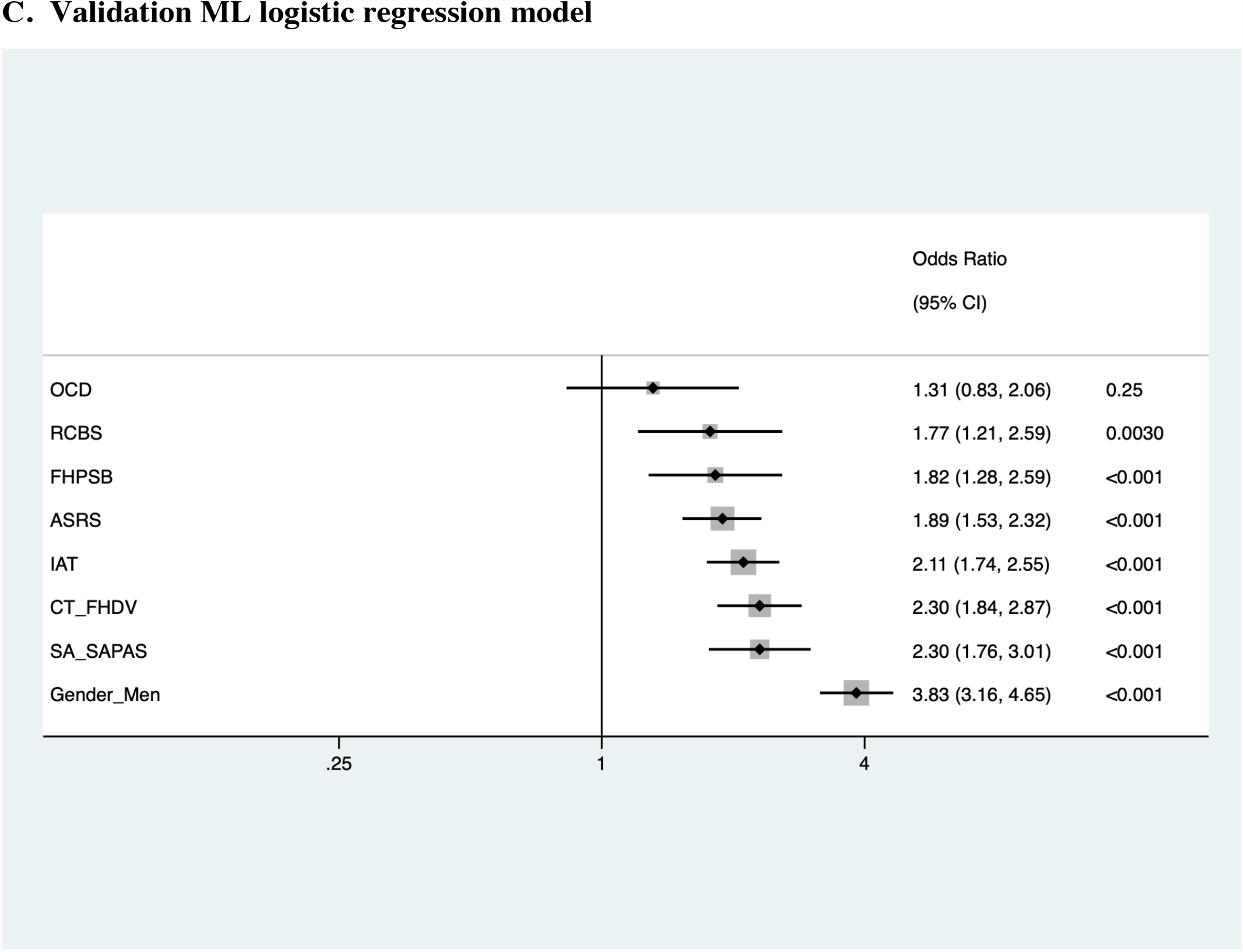
Forest plots of problematic sexual behavior (PSB) logistic regression analysis. A.The log likelihood of the model was -1149.98 (N=3049, after removing outliers, with a pseudo R^2^=0.16, *P*<0.00001). The model passed the linktest with a significant *P*(hat) (*P*<0.001) and a non-significant *P*(hatsq) (*P*=0.12). The probability of the Hosmer-Lemeshow χ^2^ test was insignificant (*P*=0.64), suggesting a good model fit. The mean variance inflation factor (VIF) and the condition number were 1.08 and 6.95, respectively. The model accuracy, area under the receiver operating (ROC) curve, sensitivity, specificity, and F score were 84.32%, 77.37%, 14.46%, 98.19% and 0.23, respectively. B. The machine learning logistic regression was conducted on the dataset without removing the outliers (training, N=2477; testing, N=619; train to test ratio: 0.80). The model accuracy, area under the ROC curve, sensitivity, specificity and F score was 74.64%.73.90%, 53.55%, 79.76% and 0.45, respectively. C. The validation model was trained by adjusted PSB, N=3141, and was tested by monthly PSB, N=3145. The model accuracy, area under the ROC curve, sensitivity, specificity and F score was 75.04%.74.26%, 59.11%, 76.97% and 0.34, respectively. OCD: obsessive-compulsive disorder (including self-reported and MINI-diagnosis); ASRS: the Adult ADHD Self-Report Scale (v1.1); IAT: the Internet Addiction Test; FHPSB: a family history of problematic sexual behavior (including a family history of sex addiction and SAST-R Core question 2); SA-SAPAS: Standardized Assessment of Personality–Abbreviated Scale as a Self-Administered Screening Test; CT_FHDV: childhood trauma and a family history of domestic violence (including physical, verbal/emotional and sexual childhood trauma and SAST-R Core question 1); RCBS: Richmond Compulsive Buying Scale

### Machine Learning (ML) logistic regression

Our ML logistic regression showed that RCBS (*P*=0.0030), SA-SAPAS (*P*<0.001), OCD (*P*<0.001), a family history of PSB (*P*<0.001), ASRS (*P*<0.001), childhood trauma (*P*<0.001), IAT (*P*<0.001), and gender (men) (*P*<0.001) were all associated with increased odds of PSB (Figure 2B). A validation of the existing model was also undertaken. The validation model was trained with SAST-R Core (subtracting SASTC1 and SASTC2, N=3141) and tested with self-reported PSB (N=3145, Figure 2C). Sensitivity analyses were performed using unadjusted SAST-R Core and family history of sexual addiction by logistic regression and ML logistic regression (Supplemental Digital Content, Figure 3-4).

**Figure 3.**
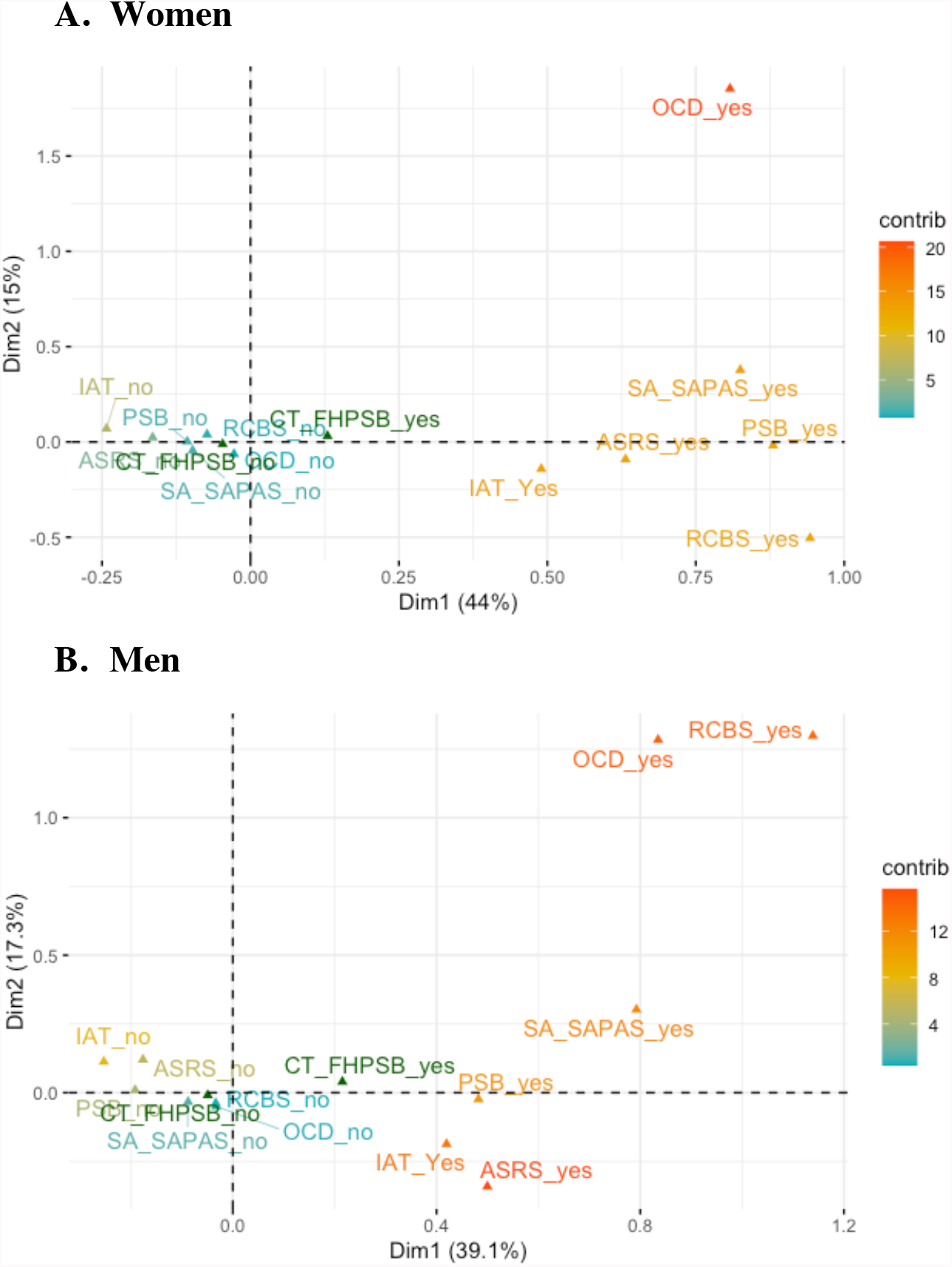
Results of multiple correspondence analysis (dimensions 1 and 2) in women (A) and men (B) The maps of data binarized into two categories (yes/no, for each variable) in dimensions 1 and 2 in women and men are shown. OCD: obsessive-compulsive disorder (including self-reported and MINI-diagnosis); SA-SAPAS: Standardized Assessment of Personality–Abbreviated Scale as a Self-Administered Screening Test; ASRS: the full Adult ADHD Self-Report Scale (v1.1); IAT: the Internet Addiction Test; RCBS: Richmond Compulsive Buying Scale; CT_FHPSB: childhood trauma and a family history of problematic sexual behavior

**Figure 4.**
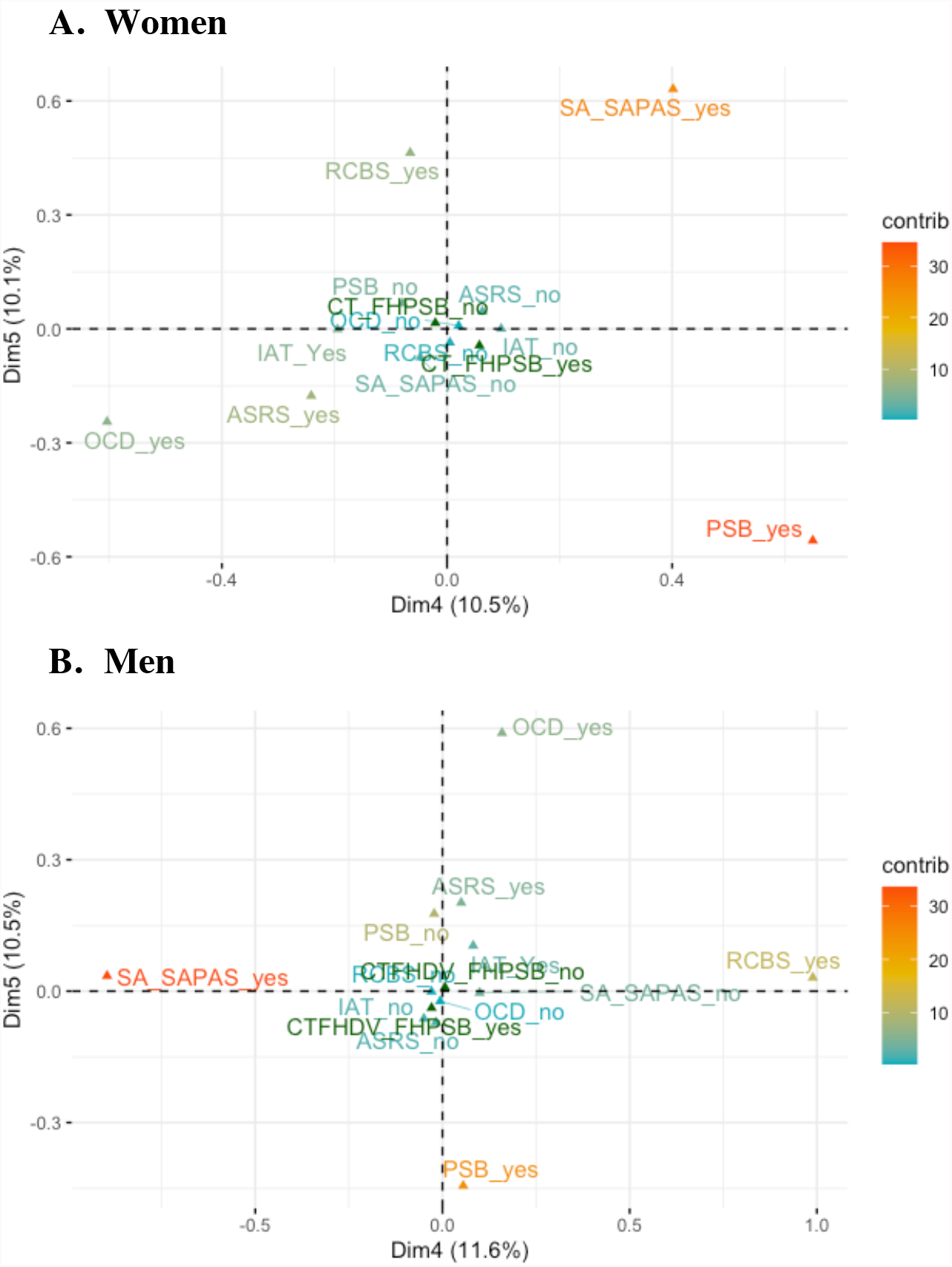
Results of multiple correspondence analysis (dimensions 4 and 5) in women (A) and men (B) The maps of data binarized into two categories (yes/no, for each variable) in dimensions 4 and 5 in women and men are shown. OCD: obsessive-compulsive disorder (including self-reported and MINI-diagnosis); SA-SAPAS: Standardized Assessment of Personality–Abbreviated Scale as a Self-Administered Screening Test; ASRS: the full Adult ADHD Self-Report Scale (v1.1); IAT: the Internet Addiction Test; RCBS: Richmond Compulsive Buying Scale; CT_FHPSB: childhood trauma and a family history of problematic sexual behavior

### Multiple correspondence analysis (MCA)

The MCA results showed that in both women and men, the first two dimensions accounted for the most explained variance (Supplemental Digital Content, Figure 5). PSB was strongly represented in both men and women in dimensions 1, 2, 4 and 5, although showing slightly different data clustering patterns by gender in these dimensions (Figure 3-4). In dimension 1, PSB clustered with all variables in women and men. In dimension 2 (Figure 3 and Supplemental Digital Content, Figure 5), PSB clustered with IAT and ASRS in men, and with RCBS, IAT and ASRS in women. In women and men, dimensions 4 and 5 illustrate that PSB was clustered relatively closely to the composite childhood trauma variable. (Figure 4, Supplemental Digital Content, Figure 5). The quality of representation and contributions of each variable are shown in Supplemental Digital Content, Table 6.

## DISCUSSION

The frequency of problematic sexual behavior, as defined by screening positive on the SAST-R Core, was 8.49% in the women and 19.7% in the men in this post-secondary education sample, consistent with prior reports in this population.^15-17^ Factors consistently associated with increased odds of PSB were gender, mental health conditions (ADHD, personality disorder, and OCD), other problematic behaviors such as internet addiction (IAT) and compulsive buying (RCBS), childhood trauma (including a family history of domestic violence), and a family history of PSB. Our results also showed that the self-reported data were correlated with the screening tool and/or MINI data (Supplemental Digital Content, Table 3).

Model validation (training with SAST-R Core subtracting the first two questions, and testing with self-reported PSB) resulted in very similar model fitting statistics (Figures 2B and 2C). A sensitivity analysis (Supplemental Digital Content, Figure 3-4) was undertaken using the original variables, namely SAST-R Core (without subtracting the first two question), childhood trauma (without adding the first question of SAST-R Core or a family history of domestic violence) and a family history of sex addiction (without incorporating SASTC2). The logistic regression model using these variables (pseudo *R*^*2*^=0.13) revealed that RCBS (*P*=0.033), ASRS (*P*<0.001), IAT (*P*<0.001), OCD (*P*<0.001), childhood trauma (*P*<0.001), and gender (baseline: women, *P*<0.001) remained significant. A family history of sex addiction was the only variable that became non-significant (*P*=0.17), probably due to the lower frequency of endorsement of this variable (N=49) compared to that of SASTC2. The combined variable (a family history of sex addiction with SASTC2, 6.55% endorsement: 214/3268), was, however, significant in all models. Using the new PSB variable, including childhood trauma and a family history of PSB but excluding sexual orientation as well as current mental health condition(s) improved the model fit slightly in the logistic regression models (e.g., sensitivity: unadjusted 77.20% vs adjusted 79.70%, F score: unadjusted 0.42 vs. adjusted 0.45). In general, the ML logistic regression models showed a higher sensitivity and F score compared to conventional logistic regression.

The variables previously associated with the RDS construct, such as SA-SAPAS, ASRS, and IAT, were associated with increased odds of PSB. In the final logistic (Figure 2) and ML logistic regressions (Figure 2), RCBS was associated with increased odds of PSB. Of note, there were gender differences regarding RCBS: RCBS was negatively correlated with men (Figure 1, baseline: women), and PSB displayed a weaker association with RCBS in men on MCA (Figure 3).

In addition, interestingly, the distribution of PSB differed significantly between OCD-positive and -negative groups (*P*<0.001, Table 1), and PSB was correlated with OCD (ϱ=0.27, *P*<0.001, Figure 1, Supplemental Digital Content, Table 3). In our final regression models and sensitivity analyses (by both conventional and ML logistic regression), OCD was associated with increased odds of PSB (OR 1.79, *P*=0.015, and OR 1.85, *P*=0.013 for conventional and ML logistic regression respectively; sensitivity models: OR 2.28, *P*<0.001, and OR 1.73, *P*=0.033, respectively).

### Limitations

Limitations of this study include the following. Firstly, as respondents to the email from the registrar knew the subject area of the study, there is a potential selection bias. However, of note, the frequency of PSB in our dataset was comparable to that seen in other studies of adults in post-secondary education as mentioned above.^12,17^ Subsample sizes were another limiting factor. Specifically, as the frequency of endorsement of OCD was relatively low (120/3206, 3.74%), although it appears on the MCA that PSB clusters more closely to the measures of reward deficiency than to OCD, this should not be overinterpreted. For sexual orientation and gender, the smaller groups were combined in the former case and dropped in the latter. For the former, bisexual (366/3371, 10.86%), homosexual (129/3371, 3.83%), and other (91/3371, 2.70%) were regrouped as “non-heterosexual sexual orientation.” Due to the small numbers in the gender minority groups (trans and other: 47/3375, 1.39%), only men and women were included in the logistic regression analysis. Another potential limitation is that we dropped missing values (rather than using imputation). However, if the proportion of missing data is relatively small (5% or less), this should not be problematic.^36^ We also dropped two measures, the FTND (0.45%, 15/3332) and PGDF (2.28%, 76/3332) due to very low endorsement rates (which were lower than in another university student sample).^37^ Although various methods may be used to impute data, some such methods (e.g., monotone imputation, or chained equations) may introduce bias, and perfect (inaccurate) predictions, as the methods themselves involve the fitting of a regression model for a categorical outcome.^38^ In our sample, the feature with the most missing data in the SAST-R Core scale was question 14, which is about feeling depressed after having sex, with only 16 out of 3341 (0.48%) endorsed (and out of those 16 participants, only 1 scored as SAST-R Core positive). Another limitation is that logistic regression is sensitive to high correlations between predictor variables. Although several variables were correlated with each other in our dataset, the correlation coefficients were less than 0.80,^39^ and the mean variance inflation factor (VIF) and the condition number were at acceptable levels (VIF<10, condition number<10) in our final models,^40^ indicating that multi-collinearity was not a significant concern. A power calculation using STATA using the parameters for OCD (the weakest predictor, P1=0.16, P2=0.24, pseudo *R*^*2*^=0.16) that for an α of 0.05, a sample size of only 264 would give a power of 0.8. Finally, the proportion of the variance (pseudo *R*^*2*^=0.16, Figure 2) explained by our analysis was relatively low. Of note, the variables that we included in our models were driven by the prior literature. Future research could include feature selection techniques, alternative machine learning models, and consideration of genetic and other biological variables. Validation through additional independent datasets with overlapping variables would also advantageous.

## CONCLUSIONS

In summary, we have identified associations between problematic sexual behavior in adults in post-secondary education and the following: compulsive buying, personality disorder, OCD, ADHD, childhood trauma, a family history of PSB, internet addiction, and male gender. Our results not only suggest that measures associated with reward deficiency syndrome are associated with PSB, but also that OCD is a vulnerability factor. Childhood trauma and a family history of domestic violence also showed associations with PSB, implying that emotional regulation and attachment style may be involved in the development of PSB. Our data are consistent with different subgroups within PSB with slightly somewhat contrasting etiological mechanisms by gender.

## Data Availability

Data produced in the present study are available in deidentified format upon reasonable request to the authors.

## Acknowledgments

We would like to acknowledge the contribution of Dr. Patrick Carnes to the study design, and volunteer input of Penny Carnes, Grace Li, Hana Graham, and Chanelle Martens to data collection. SJ would like to thank Afia Anjum from DW’s lab for her assistance with R. There was no pharmaceutical or industry support for the work reported herein.

