## Supplemental Material for "Logistic regression with machine learning sheds light on the problematic sexual behavior phenotype"

**Supplemental Table 1. Endorsement rates of SAST-R Core questions, with rates in variables defining problematic sexual behaviour (PSB) in different manners**

| **Number** | **Question** | **Endorsement** | **“Yes” vs. self-reported monthly PSB** | **“Yes” vs. SAST-R Core**  **(unadjusted scores)** | **“Yes” vs. SAST-R Core (adjusted)** | **“Yes” vs. PSB (adjusted)** |
| --- | --- | --- | --- | --- | --- | --- |
| 1. | SASTC11 -- Hide some of your sexual behaviors from others  (Affect disturbance) | 1234/3341  (36.94%) | (1)  255/350  (72.86%) | (1)  362/406  (89.16%) | (1)  330/359  (91.92%) | (1)  419/532  (78.76%) |
| 2. | SASTC3 -- Find yourself preoccupied with sexual thoughts  (Preoccupation) | 805/3335 (24.14%) | (4)  194/348 (55.75%) | (4)  290/406  (71.43%) | (4)  261/359  (72.70%) | (3)  330/532  (62.03%) |
| 3. | SASTC19 -- Sex been a way to escape your problems  (Preoccupation) | 780/3340  (23.35%) | (5)  191/350  (54.57%) | (5)  277/406  (68.23%) | (5)  243/359  (67.69%) | (5)  311/532  (58.46%) |
| 4. | SASTC5 -- Feel bad about your sexual behavior  (Affect disturbance) | 718/3339 (21.50%) | (3)  195/349  (55.87%) | (3)  305/406  (75.12%) | (3)  277/359 (77.16%) | (4)  327/532  (61.47%) |
| 5. | SASTC12 -- Attempted to stop some parts of your sexual activity  (Loss of control) | 664/3337 (19.9%) | (2)  209/350  (59.71%) | (2)  319/406  (78.57%) | (2)  289/359  (80.50%) | (2)  350/532  (65.79%) |
| 6. | SASTC13 -- Felt degraded by your sexual behaviors  (Affect disturbance) | 478/3339 (14.32%) | (7)  137/350  (39.14%) | (6)  260/407  (63.88%) | (6)  241/359  (67.13%) | (6)  261/532  (49.06%) |
| 7. | SASTC10 -- Made efforts to quit a type of sexual activity and failed  (Loss of control) | 398/3340 (11.92%) | (6)  155/349  (44.41%) | (7)  233/407  (57.25%) | (7)  218/359  (60.72%) | (7)  254/532  (47.74%) |
| 8. | SASTC4 -- Feel that your sexual behavior is not normal  (Affect disturbance) | 371/3342 (11.10%) | 106/350  (30.29%) | 167/406  (41.13%) | 151/359 (42.06%) | 177/532  (33.27%) |
| 9. | SASTC1 -- Sexually abused as a child or adolescent  (Associated features) | 367/3344 (10.97%) | 60/350 (17.14%) | 110/407  (27.03%) | 72/359  (20.06%) | 96/532  (18.05%) |
| 10. | SASTC8 -- Anyone hurt emotionally because of your sexual behavior  (Relationship disturbance) | 310/3343 (9.27%) | 73/350  (20.86%) | 144/407  (35.38%) | 135/359  (37.60%) | 151/532  (28.38%) |
| 11. | SASTC15 -- Feel controlled by your sexual desire  (Loss of control) | 231/3340 (6.92%) | 93/350  (26.57%) | 144/407  (35.38%) | 138/359  (38.44%) | 156/532  (29.32%) |
| 12. | SASTC14 – Feel depressed after having Sex  (Affect disturbance) | 201/3331 (6.03%) | 55/350  (15.71%) | 99/406  (24.38%) | 89/359  (24.79%) | 100/532  (18.80%) |
| 13. | SASTC17 -- Think your sexual desire is stronger than you are  (Loss of control) | 185/3335 (5.55%) | 80/349  (22.92%) | 124/404  (30.69%) | 120/359  (33.43%) | 135/532  (25.38%) |
| 14. | SASTC2 -- Parents have trouble with sexual behavior  (Associated features) | 184/3340 (5.51%) | 39/350 (11.14%) | 71/407  (17.44%) | 48/359  (13.37%) | 63/532  (11.84%) |
| 15. | SASTC6 -- Sexual behavior created problems for you and family  (Relationship disturbance) | 180/3343 (5.38%) | 62/350  (17.71%) | 110/407  (27.03%) | 106/359  (29.53%) | 112/532  (21.05%) |
| 16. | SASTC16 -- Parts of your life neglected because spending too much time on Sex  (Relationship disturbance) | 136/3338 (4.07%) | 66/349  (18.91%) | 96/405  (23.70%) | 89/359  (24.79%) | 106/532  (19.92%) |
| 17. | SASTC7 -- Sought help for sexual behavior  (Associated features) | 90/3340 (2.69%) | 33/350  (9.43%) | 64/407  (15.72%) | 60/359  (16.71%) | 62/532  (11.65%) |
| 18. | SASTC18 -- Sex almost all you think about  (Preoccupation) | 69/3341  (2.07%) | 24/350  (6.86%) | 46/407  (11.30%) | 43/359  (11.98%) | 45/532  (8.46%) |
| 19. | SASTC20 -- Sex most important thing in your life  (Preoccupation) | 37/3340 (1.11%) | 15/349  (4.30%) | 25/405  (6.17%) | 25/359  (6.96%) | 25/532  (4.70%) |
| 20. | SASTC9 -- Sexual activities against the law  (Associated features) | 35/3342 (1.05%) | 8/349  (2.29%) | 24/405  (5.93%) | 21/359  (5.85%) | 22/532  (4.14%) |

The top six endorsements come from the affect disturbance, preoccupation and loss of control categories in the SAST-R Core. The ranking of top endorsements by monthly PSB and SAST-R Core were similar, except for SASTC13 and SASTC10. Subtracting SASTC1 and SASTC2 and combining monthly PSB did not change the top six endorsements.

SASTC: SAST-R Core question

**Supplemental Table 2. Distribution of gender and sexual orientation by those screening positive on the unadjusted SAST-R Core in participants**

| **Gender** | **SAST-R Core** | | **P value** |
| --- | --- | --- | --- |
|  | **Yes** | **No** |  |
| Women  (N=2226) | 189 (8.49%) | 2037 (91.51%) | *P*<0.001^a^ |
| Men  (N=1066) | 210 (19.70%) | 856 (80.30%) |  |
| Total number  (N=3292) | 399 (12.12%) | 2893 (87.88%) |  |
| **Sexual Orientation** | **SAST-R Core** | | *P*<0.001^a^ |
|  | **Yes** | **No** |  |
| Heterosexual  (N=2754) | 308 (11.18%) | 2446 (88.82%) |  |
| Non-heterosexual  (N=581) | 98 (16.87%) | 483 (83.13%) |  |
| Total number (N=3335) | 406 (12.17%) | 2929 (87.83%) |  |

^a^Pearson χ^2^ test

**Supplemental Table 3. Correlation matrix between problematic sexual behavior (PSB) with demographic and clinical variables**

| **Tetrachoric ρ,**  ***P*** | **PSB** | **Non_heterosexual** | **MH_condition** | **CT_FHDV** | **RCBS** | **OCD** | **SA-SAPAS** | **FHPSB** | **ASRS** | **IAT** |
| --- | --- | --- | --- | --- | --- | --- | --- | --- | --- | --- |
| **Non_heterosexual** | **0.084**  **(*P*=0.036)** |  |  |  |  |  |  |  |  |  |
| **MH_condition** | **0.12**  **(*P*=0.0013)** | **0.32**  **(*P*<0.001)** |  |  |  |  |  |  |  |  |
| **CT_FHDV** | **0.24**  **(*P*<0.001)** | **0.23**  **(*P*<0.001)** | **0.37**  **(*P*<0.001)** |  |  |  |  |  |  |  |
| **RCBS** | **0.27**  **(*P*<0.001)** | -0.050  (*P*=0.41) | **0.16**  **(*P*=0.0013)** | **0.15**  **(*P*=0.0017)** |  |  |  |  |  |  |
| **OCD** | **0.27**  **(*P*<0.001)** | **0.17**  **(*P*=0.0041)** | **0.42**  **(*P*<0.001)** | **0.21**  **(*P*<0.001)** | **0.19**  **(*P*=0.0094)** |  |  |  |  |  |
| **SA-SAPAS** | **0.30**  **(*P*<0.001)** | **0.14**  **(*P*=0.0010)** | **0.27**  **(*P*<0.001)** | **0.19**  **(*P*<0.001)** | **0.26**  **(*P*<0.001)** | **0.31**  **(*P*<0.001)** |  |  |  |  |
| **FHPSB** | **0.30**  **(*P*<0.001)** | **0.11**  **(*P*=0.022)** | **0.20**  **(*P*<0.001)** | **0.69**  **(*P*<0.001)** | 0.074  (*P*=0.27) | **0.19**  **(*P*=0.0079)** | **0.13**  **(*P*=0.0152)** |  |  |  |
| **ASRS** | **0.33**  **(*P*<0.001)** | **0.21**  **(*P*<0.001)** | **0.27**  **(*P*<0.001)** | **0.13**  **(*P*<0.001)** | **0.21**  **(*P*<0.001)** | **0.15**  **(*P*=0.0051)** | **0.31**  **(*P*<0.001)** | 0.081  (*P*=0.093) |  |  |
| **IAT** | **0.34**  **(*P*<0.001)** | **0.098**  **(*P*=0.0040)** | **0.13**  **(*P*<0.001)** | **0.071**  **(*P*=0.027)** | **0.40**  **(*P*<0.001)** | **0.15**  **(*P*=0.0051)** | **0.31**  **(*P*<0.001)** | 0.061  (*P*=0.19) | **0.43**  **(*P*<0.001)** |  |
| **Gender_men** | **0.40**  **(*P*<0.001)** | **-0.12**  **(*P*=0.0013)** | **-0.26**  **(*P*<0.001)** | **-0.17**  **(*P*<0.001)** | **-0.26**  **(*P*<0.001)** | 0.035  (*P*=0.53) | 0.31  (*P*=0.66) | 0.011  (*P*=0.81) | **0.11**  **(*P*=0.0011)** | **0.075**  **(*P*=0.014)** |

MH_condition: mental health condition; CT_FHDV: childhood trauma and a family history of domestic violence (including physical, verbal/emotional and sexual childhood trauma and SAST Core question 1); RCBS: Richmond Compulsive Buying Scale; OCD: obsessive-compulsive disorder (including self-reported and MINI-diagnosis); SA-SAPAS: Standardized Assessment of Personality–Abbreviated Scale as a Self-Administered Screening Test; FHPSB: a family history of problematic sexual behavior (including a family history of sexual addiction and SAST Core question 2); ASRS: the Adult ADHD Self-Report Scale (v1.1); IAT: the Internet Addiction Test

**Supplemental Table 4. Correlation matrices between derived and original variables**

A. **Between (adjusted) problematic sexual behavior (PSB), SAST-R Core (unadjusted), and self-reported PSB in the past 30 days**

| **Tetrachoric ρ,**  ***P*** | **PSB** | **SAST-R Core** | **Self-reported PSB in the past 30 days** |
| --- | --- | --- | --- |
| **PSB** | 1  (N=3219) |  |  |
| **SAST-R Core** | 0.97  (N=3219, *P*<0.001) | 1  (N=3341) |  |
| **Self-reported PSB in the past 30 days** | 1  (N=3219, *P*<0.001) | 0.70  (N=3271, *P*<0.001) | 1  (N=3271) |

B. **Between total obsessive-compulsive disorder (OCD), OCD by MINI diagnosis, and self-reported OCD**

| **Tetrachoric ρ,**  ***P*** | **OCD** | **OCD MINI diagnosis** | **Self-reported OCD** |
| --- | --- | --- | --- |
| **OCD** | 1  (N=3372) |  |  |
| **OCD MINI diagnosis** | 1  (N=1809, *P*<0.001) | 1  (N=1809) |  |
| **Self-reported OCD** | 1  (N=3348, *P*<0.001) | 0.47  (N=1785, *P*<0.001) | 1  (N=3348) |

The binary SAST-R Core variable was significantly correlated with self-reported “maximum length of time on any single occasion spent in problematic sexual activities” (τ_b_=0.32, *P*<0.001, N=2049). IAT was significantly associated with self-reported “problematic internet use in the past 30 days” (for the four severity categories of IAT, and the binary IAT variable: τ_b_=0.42, *P*< 0.001 and τ_b_=0.40, *P*<0.001, N=1441, respectively). In addition, SA-SAPAS was significantly correlated with antisocial personality disorder diagnosed by MINI (tetrachoric ρ=0.24, *P*=0.026, N=1788) and with self-reported borderline personality disorder (tetrachoric ρ=0.44, *P*<0.001, N=1429).

**Supplemental Table 5. Multiple correspondence analysis (MCA) of problematic sexual behavior (PSB): quality of representation analysis by total cosine squared (cos^2^)**

A. **Women**

| **Variable** | **Dimension 1** | **Dimension 2** | **Dimension 3** | **Dimension 4** | **Dimension 5** | **Total cos^2^** |
| --- | --- | --- | --- | --- | --- | --- |
| PSB | 0.50 | 0.00020 | 0.020 | 0.27 | 0.20 | 1.00 |
| IAT | 0.61 | 0.051 | 0.0039 | 0.096 | 0 | 0.76 |
| ASRS | 0.55 | 0.011 | 0.16 | 0.080 | 0.043 | 0.84 |
| RCBS | 0.38 | 0.11 | 0.37 | 0.0018 | 0.092 | 0.95 |
| SA-SAPAS | 0.44 | 0.092 | 0.11 | 0.10 | 0.26 | 1.00 |
| OCD | 0.13 | 0.69 | 0.098 | 0.073 | 0.012 | 1.00 |

B. **Men**

| **Variable** | **Dimension 1** | **Dimension 2** | **Dimension 3** | **Dimension 4** | **Dimension 5** | **Total cos^2^** |
| --- | --- | --- | --- | --- | --- | --- |
| PSB | 0.51 | 0.0012 | 0.058 | 0.0067 | 0.43 | 1.00 |
| IAT | 0.57 | 0.11 | 0.011 | 0.022 | 0.035 | 0.75 |
| ASRS | 0.48 | 0.22 | 0.0094 | 0.0048 | 0.078 | 0.79 |
| RCBS | 0.22 | 0.29 | 0.31 | 0.17 | 0.00020 | 0.99 |
| OCD | 0.16 | 0.37 | 0.38 | 0.0057 | 0.079 | 0.99 |
| SA-SAPAS | 0.39 | 0.057 | 0.054 | 0.50 | 0.00080 | 1.00 |

The cosine squared of each variable indicated a good representation of PSB (total cos^2^(PSB)=1) across the five dimensions in both women and men.

IAT: the Internet Addiction Test; ASRS: the Adult ADHD Self-Report Scale (v1.1); RCBS: Richmond Compulsive Buying Scale; OCD: obsessive-compulsive disorder (including self-reported and MINI-diagnosis); SA-SAPAS: Standardized Assessment of Personality–Abbreviated Scale as a Self-Administered Screening Test

**Supplemental Table 6. The contributions of each variable in dimensions 1 to 5 to the MCA**

A. **Women**

| **Variable** | **Contribution to MCA** | | | | | **Total** |
| --- | --- | --- | --- | --- | --- | --- |
|  | **Dimension 1** | **Dimension 2** | **Dimension 3** | **Dimension 4** | **Dimension 5** |  |
| PSB_no | 2.07 | 0.0026 | 0.29 | 4.72 | 3.64 | 10.72 |
| PSB_yes | 17.14 | 0.022 | 2.44 | 39.02 | 30.07 | 88.69 |
| IAT_no | 8.05 | 1.97 | 0.18 | 5.30 | 0.00020 | 15.50 |
| IAT_yes | 16.27 | 3.97 | 0.37 | 10.70 | 0.00040 | 31.31 |
| ASRS_no | 4.41 | 0.27 | 4.47 | 2.69 | 1.50 | 13.34 |
| ASRS_yes | 16.91 | 1.03 | 17.13 | 10.30 | 5.75 | 51.12 |
| SA_SAPAS_no | 1.74 | 1.07 | 1.57 | 1.72 | 4.46 | 10.56 |
| SA_SAPAS_yes | 4.72 | 9.06 | 13.28 | 14.57 | 37.78 | 79.41 |
| RCBS_no | 1.02 | 0.85 | 3.46 | 0.021 | 1.08 | 6.431 |
| RCBS_yes | 13.10 | 11.02 | 44.65 | 0.26 | 13.88 | 82.91 |
| OCD_no | 0.15 | 2.34 | 0.40 | 0.35 | 0.061 | 3.301 |
| OCD_yes | 4.42 | 68.40 | 11.75 | 10.34 | 1.77 | 96.68 |

B. **Men**

| **Variable** | **Contribution to MCA** | | | | | **Total** |
| --- | --- | --- | --- | --- | --- | --- |
|  | **Dimension 1** | **Dimension 2** | **Dimension 3** | **Dimension 4** | **Dimension 5** |  |
| PSB_no | 6.24 | 0.034 | 2.10 | 0.28 | 19.75 | 28.40 |
| PSB_yes | 15.69 | 0.084 | 5.29 | 0.70 | 49.64 | 71.40 |
| IAT_no | 9.48 | 4.26 | 0.54 | 1.23 | 2.16 | 17.67 |
| IAT_yes | 15.70 | 7.05 | 0.89 | 2.03 | 3.58 | 29.25 |
| ASRS_no | 5.45 | 5.74 | 0.32 | 0.18 | 3.32 | 15.01 |
| ASRS_yes | 15.43 | 16.28 | 0.90 | 0.52 | 9.41 | 42.54 |
| SA-SAPAS_no | 1.65 | 0.54 | 0.67 | 7.10 | 0.012 | 9.97 |
| SA-SAPAS_yes | 14.85 | 4.89 | 6.07 | 64.07 | 0.11 | 90.00 |
| RCBS_no | 0.26 | 0.76 | 1.07 | 0.66 | 0.00070 | 2.75 |
| RCBS_yes | 8.801 | 25.89 | 36.33 | 22.43 | 0.024 | 93.48 |
| OCD_no | 0.24 | 1.29 | 1.72 | 0.030 | 0.45 | 3.73 |
| OCD_yes | 6.20 | 33.17 | 44.09 | 0.76 | 11.53 | 95.75 |

PSB: problematic sexual behavior; IAT: the Internet Addiction Test; ASRS: the Adult ADHD Self-Report Scale (v1.1); RCBS: Richmond Compulsive Buying Scale; OCD: obsessive-compulsive disorder (including self-reported and MINI-diagnosis); SA-SAPAS: Standardized Assessment of Personality–Abbreviated Scale as a Self-Administered Screening Test

**Supplemental Figure 1. Study recruitment and analysis flow chart for the conventional logistic regression and multiple correspondence analysis**

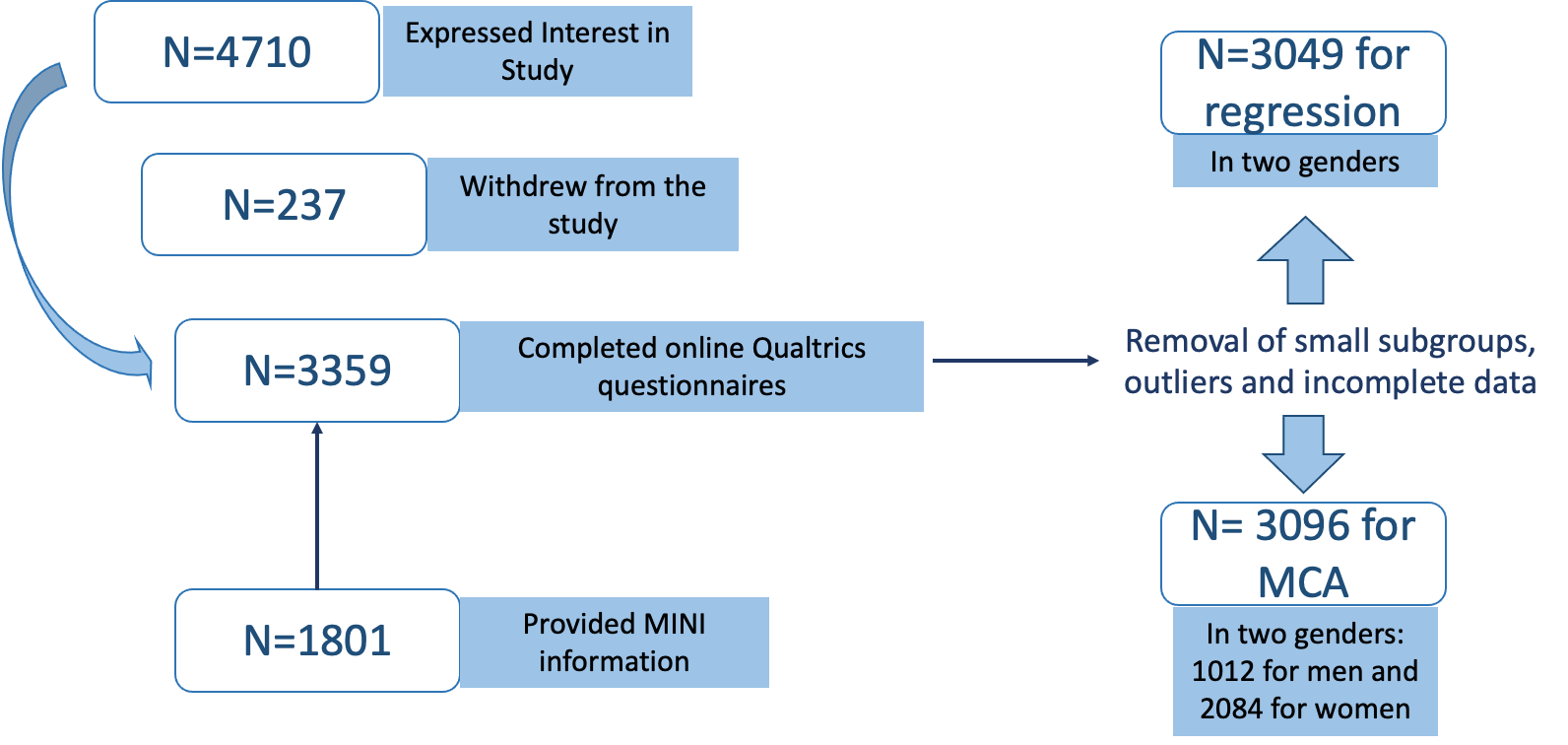

MCA: Multiple correspondence analysis; to maximize power, outliers were not removed for the MCA and the machine learning logistic regression

**Supplemental Figure 2. Figures indicating satisfactory removal of outliers for the conventional logistic regression**

A. **Standardized Pearson residual plot**

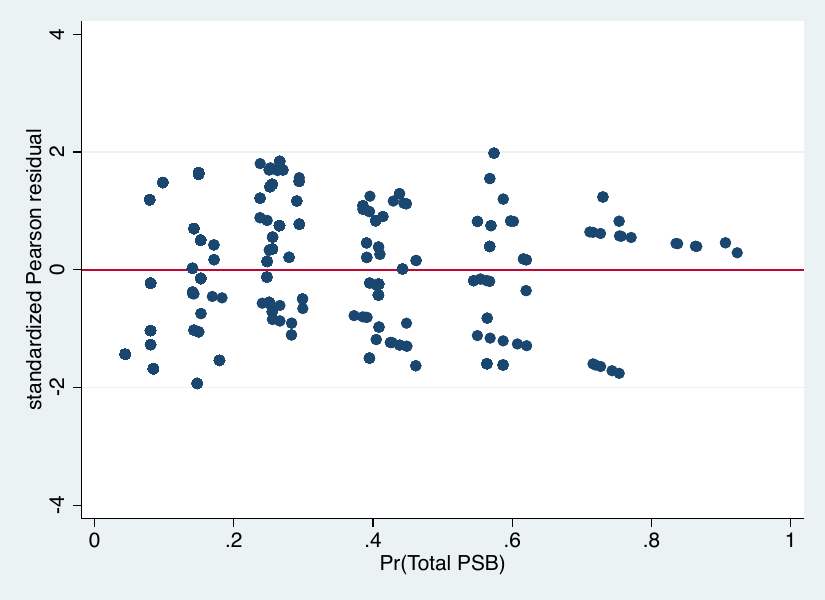

Standardized Pearson residuals were between -2 and 2 after removing the outliers.

B. **Influential points by ∆**$\hat{\boldsymbol{\beta}}$ **(dBeta)**

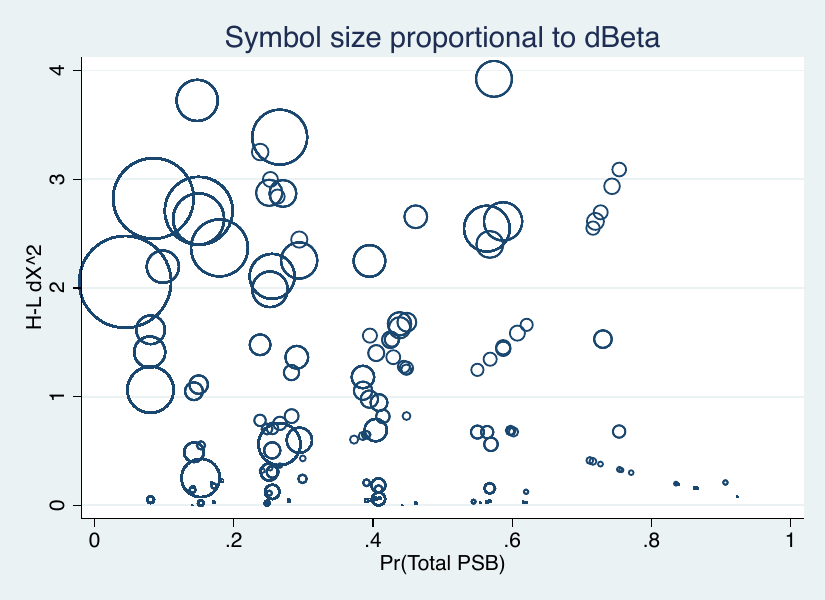

The Hosmer-Lemeshow ∆$\hat{\beta}$ was less than 4 after removing the outliers.

C. **Leverage Plot**

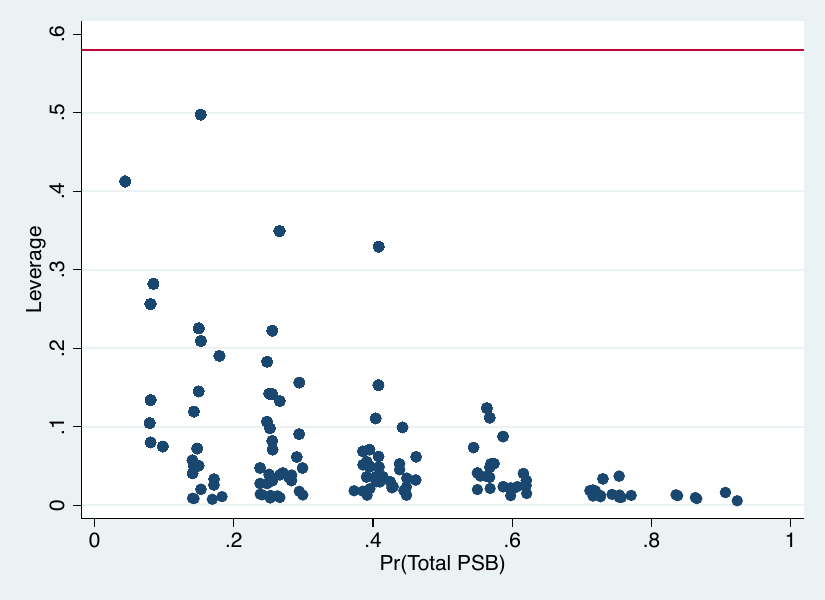

The values of leverage from the variables were lower than two times the mean (mean value 0.29).

**Supplemental Figure 3. Forest plot of logistic regression analysis with SAST-R Core (screening positive on unadjusted SAST-R Core): sensitivity analysis**

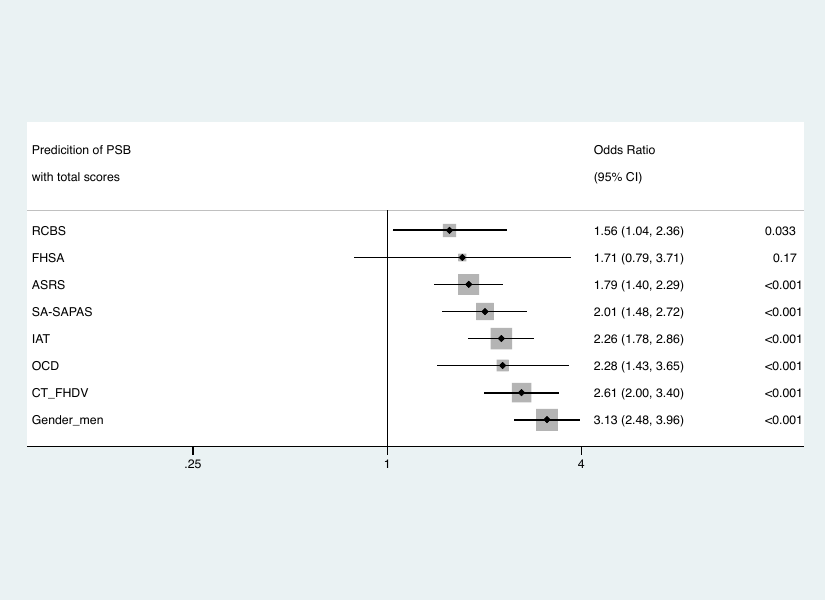

The loglikelihood of this model was -1016.19, with pseudo R^2^=0.13 (N=3186, *P*<0.001). The model passed the linktest with a significant *P*(hat) (*P*<0.001) and a non-significant *P*(hatsq) (*P*=0.71). The probability of the Hosmer-Lemeshow χ^2^ test was not significant (*P*=0.61), suggesting a good model fit. The model also passed collinearity diagnostics (mean variance inflation factor (VIF)1.05, condition number 3.11). The model accuracy, area under the receiver operating characteristic (ROC) curve, sensitivity, specificity, and F score were: 88.14%, 75.50%, 7.55%, 99.18% and 0.13, respectively.

RCBS: Richmond Compulsive Buying Scale; FHSA: a family history of sex addiction; ASRS: the Adult ADHD Self-Report Scale (v1.1); SA-SAPAS: Standardized Assessment of Personality–Abbreviated Scale as a Self-Administered Screening Test; IAT: the Internet Addiction Test; OCD: obsessive-compulsive disorder (including self-reported and MINI diagnosis); CT_FHDV: childhood trauma and a family history of domestic violence (including physical, verbal/emotional and sexual childhood trauma)

**Supplemental Figure 4. Forest plot of machine learning logistic regression analysis with SAST-R Core (screening positive on unadjusted SAST-R Core): sensitivity analysis**

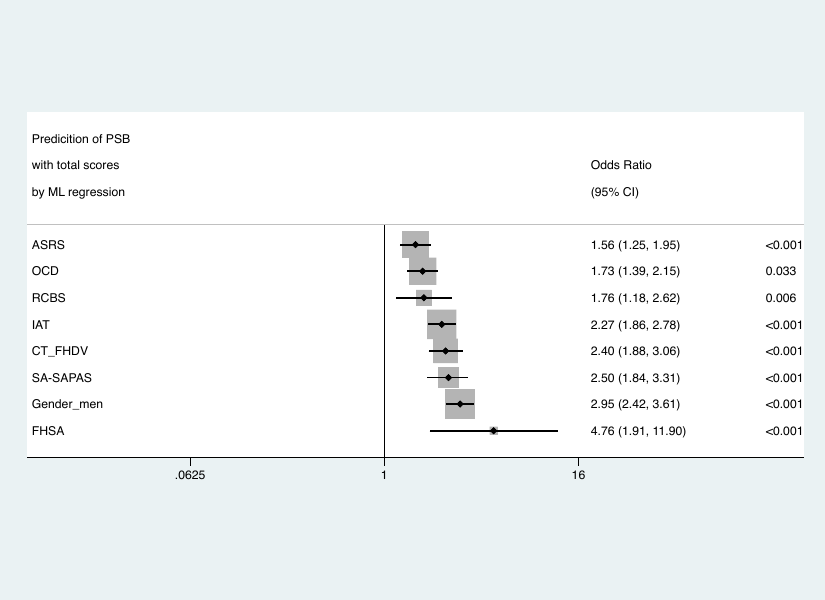

The model accuracy, area under the ROC curve, sensitivity, specificity, and F score were: 76.68%, 77.20%, 63.95%, 78.66% and 0.42, respectively (N=3186).

ASRS: the Adult ADHD Self-Report Scale (v1.1); OCD: obsessive-compulsive disorder (including self-reported and MINI diagnosis); RCBS: Richmond Compulsive Buying Scale; IAT: the Internet Addiction Test; CT_FHDV: childhood trauma and a family history of domestic violence (including physical, verbal/emotional and sexual childhood trauma); SA-SAPAS: Standardized Assessment of Personality–Abbreviated Scale as a Self-Administered Screening Test; FHSA: a family history of sex addiction

**Supplemental Figure 5. Scree plot of multiple correspondence analysis (MCA) in women (A) and men (B)**

A. **Women**

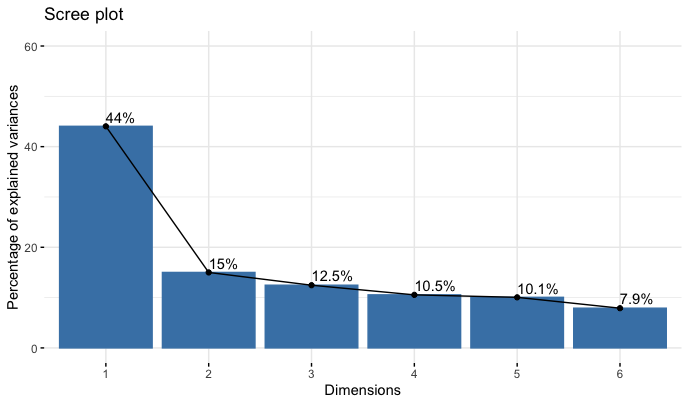

B. **Men**

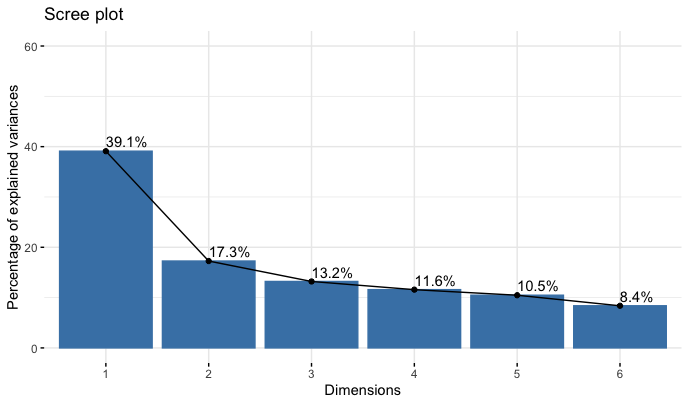
